# Association between complement component 4A expression, cognitive performance and brain imaging measures in UK Biobank

**DOI:** 10.1101/2020.06.05.20122978

**Authors:** Kevin S O’Connell, Ida E. Sønderby, Oleksandr Frei, Dennis van der Meer, Lavinia Athanasiu, Olav B. Smeland, Dag Alnæs, Tobias Kaufmann, Lars T. Westlye, Vidar M. Steen, Ole A. Andreassen, Timothy Hughes, Srdjan Djurovic

**Author notes:** These authors have equal contribution (Timothy Hughes, Srdjan Djurovic).

## Abstract

Altered expression of the complement component *C4A* gene is a known risk factor for schizophrenia. Further, predicted brain *C4A* expression has also been associated with memory function highlighting that altered C4A expression in the brain may be relevant for cognitive and behavioral traits. We investigated if predicted brain *C4A* expression was associated with cognitive performance and brain imaging measures in the UK Biobank. We identified significant negative associations between predicted *C4A* expression and performance on select cognitive tests, and significant associations with MRI-based cortical thickness and surface area in select regions. Finally, we observed significant inconsistent partial mediation of the effects of predicted *C4A* expression on cognitive performance, by specific brain structure measures. These results demonstrate that the *C4* risk locus is associated with the central endophenotypes of cognitive performance and brain morphology, even when considered independently of other genetic risk factors and in individuals without mental or neurological disorders.

## INTRODUCTION

The major histocompatibility complex (MHC) is located on chromosome 6 and is implicated in a number of autoimmune diseases (Howson et al., 2009; Kamitaki et al., 2020; Raychaudhuri et al., 2012). In addition, genetic variants within this region are consistently associated with risk of schizophrenia (International Schizophrenia Consortium et al., 2009; Pardiñas et al., 2018; Schizophrenia Psychiatric Genome-Wide Association Study (GWAS) Consortium, 2011; Schizophrenia Working Group of the Psychiatric Genomics Consortium, 2014; J. Shi et al., 2009; Stefansson et al., 2009), but not depression (Glanville et al., 2019). These associations corroborate serological studies which identified altered levels of inflammatory markers in schizophrenia patients, including complement proteins (Hakobyan et al., 2005; Laskaris et al., 2019; Maes et al., 1997; Mayilyan, Dodds, et al., 2008; Mayilyan et al., 2006; Mayilyan, Weinberger, et al., 2008). These findings suggest the involvement of an immune component in psychiatric disorders such as schizophrenia.

In order to better understand the mechanisms underlying the MHC genetic association with schizophrenia, Sekar et al. (Sekar et al., 2016) conducted a fine-mapping molecular investigation of the region and identified that variants within the complement component 4 (*C4*) gene locus are responsible for at least part of the association signal. The C4 protein is one of a number of proteins that makes up the complement system (Charles A Janeway et al., 2001), part of the innate immune system. Complement components were initially shown to modulate neurogenesis in murine primary cortical cell cultures (van Beek et al., 2001). Further investigation of the role of complement components in the central nervous system of genetically modified mice identified its major role in modulating synaptic plasticity (Hong et al., 2016; Q. Shi et al., 2015; Stephan et al., 2012; Stokowska et al., 2017; Vasek et al., 2016). More recently, complement components were implicated in neuronal migration (Gorelik et al., 2017) and apoptosis (Niculescu et al., 2004) in the central nervous system. Additional evidence for the activity of the complement system in the brain, and its involvement in the pathogenesis of schizophrenia is summarized in recent reviews (Druart & Le Magueresse, 2019; Nimgaonkar et al., 2017; Tenner et al., 2018; Woo et al., 2019).

The *C4* gene is present as one of two isotypes (*C4A* and *C4B*) and the structural variation between these isotypes, as well as their copy number, was shown to significantly alter the expression level of C4 in post-mortem brain tissue (Sekar et al., 2016). A model of this relationship can be used to predict *C4A* gene expression in the brain based on an individual’s genotype (Sekar et al., 2016). Using this procedure, predicted *C4A* gene expression was associated with risk of schizophrenia in an independent sample (Sekar et al., 2016). Finally, C4 proteins localized to the synapses in post-mortem human brains, and C4 was also demonstrated to modulate synaptic pruning in mice (Sekar et al., 2016), and human-derived neural cultures (C. M. Sellgren et al., 2017; Carl M. Sellgren et al., 2019).

Independent of these findings, variants within the MHC region were also associated with cognitive performance (Athanasiu et al., 2017; Donohoe et al., 2013; Zhang et al., 2017) and brain structure (Walters et al., 2013) in patients with schizophrenia. Based on these studies, Donohoe and colleagues (Donohoe et al., 2018) showed that increased predicted *C4A* expression was associated with poorer performance in memory recall measures in a cohort of psychosis patients and healthy controls (n = 1,238), as well as in patients only (Donohoe et al., 2018). The direction of effect in control participants was similar to that observed in patients, however the effect size was smaller and non-significant (Donohoe et al., 2018). In addition, they demonstrated that higher predicted *C4A* expression was associated with lower cortical activity in the middle temporal cortex during visual processing in healthy participants (Donohoe et al., 2018). In support of these findings, complement-dependent synapse elimination was recently identified as a mechanism for memory loss (Wang et al., 2020). These results highlight that *C4A* expression in the brain may be associated cognitive and behavioral traits not only in patients with psychiatric disorders, but also in healthy individuals.

Based on this, our primary aim was to investigate if predicted brain *C4A* expression is associated with cognitive performance in a large adult population-based sample (UK Biobank, (Bycroft et al., 2018; Miller et al., 2016)), without mental or neurological disorders. We hypothesized that higher predicted *C4A* expression would be associated with lower cognitive performance. Our secondary aims were to investigate if predicted brain *C4A* expression is associated with differences in brain structure, and if observed effects on cognitive performance may be mediated by C4A-associated differences in brain structure.

## RESULTS

### Effect of *C4A* expression on Cognitive Performance

Predicted *C4A* expression was significantly (FDR < 0.05) associated with three of the seven cognitive tests (Figure 1i, Table 1, Supplementary Table 1). Specifically, higher predicted *C4A* expression was associated with reduced cognitive performance in the pairs matching (FDR = 0.009), fluid intelligence (FDR = 0.032) and symbol digit substitution (FDR = 0.043) cognitive tasks. Analysis of the association between predicted *C4A* expression and cognitive performance measures indicates a linear relationship, not a distinct range of expression above or below which the observed changes occur (Supplementary Tables 2–4, No significant *C4A*-sex interactions were identified for any of the cognitive tests (Supplementary Table 5). Higher predicted *C4A* expression was associated with reduced cognitive performance in the pairs matching task in females only (Std. β = –0.006, FDR = 0.018) (Supplementary Table 6), but was not significant in males only despite a similar effect size (Std. β = –0.006, FDR = 0.177) (Supplementary Table 7).

**Table 1.** A summary of the results from the significant linear regression models of predicted *C4A*

| Phenotype and Covariates | Std. $\beta$ | Std. Error | Uncorrected p-value | FDR |
| --- | --- | --- | --- | --- |
| **Pairs matching** |  |  |  |  |
| *C4A* expression | -0.006 | 0.003 | $1.046 \times 10^{-3}$ | $9.212 \times 10^{-3}$ |
| Age | -0.135 | 0.675 | $< 1 \times 10^{-300}$ | $< 1 \times 10^{-300}$ |
| Sex (Male) | 0.016 | 0.002 | $4.204 \times 10^{-20}$ | $5.028 \times 10^{-19}$ |
| **Fluid Intelligence** |  |  |  |  |
| *C4A* expression | -0.008 | 0.016 | $4.174 \times 10^{-3}$ | 0.032 |
| Age | -0.011 | 0.001 | $2.415 \times 10^{-4}$ | $2.319 \times 10^{-3}$ |
| Sex (Male) | 0.059 | 0.012 | $1.209 \times 10^{-101}$ | $2.838 \times 10^{-100}$ |
| **Symbol Digit Substitution** |  |  |  |  |
| *C4A* expression | -0.008 | 0.040 | $5.930 \times 10^{-3}$ | 0.043 |
| Age | -0.455 | 0.002 | $< 1 \times 10^{-300}$ | $< 1 \times 10^{-300}$ |
| Sex (Male) | -0.006 | 0.029 | $5.477 \times 10^{-2}$ | 0.248 |

All models also included age squared, educational attainment, genotyping batch and the first 10 genetic principal components as covariates (data not shown). Std. $\beta$ = Standardized $\beta$ . Std. Error = Standard Error.

**Figure 1.**
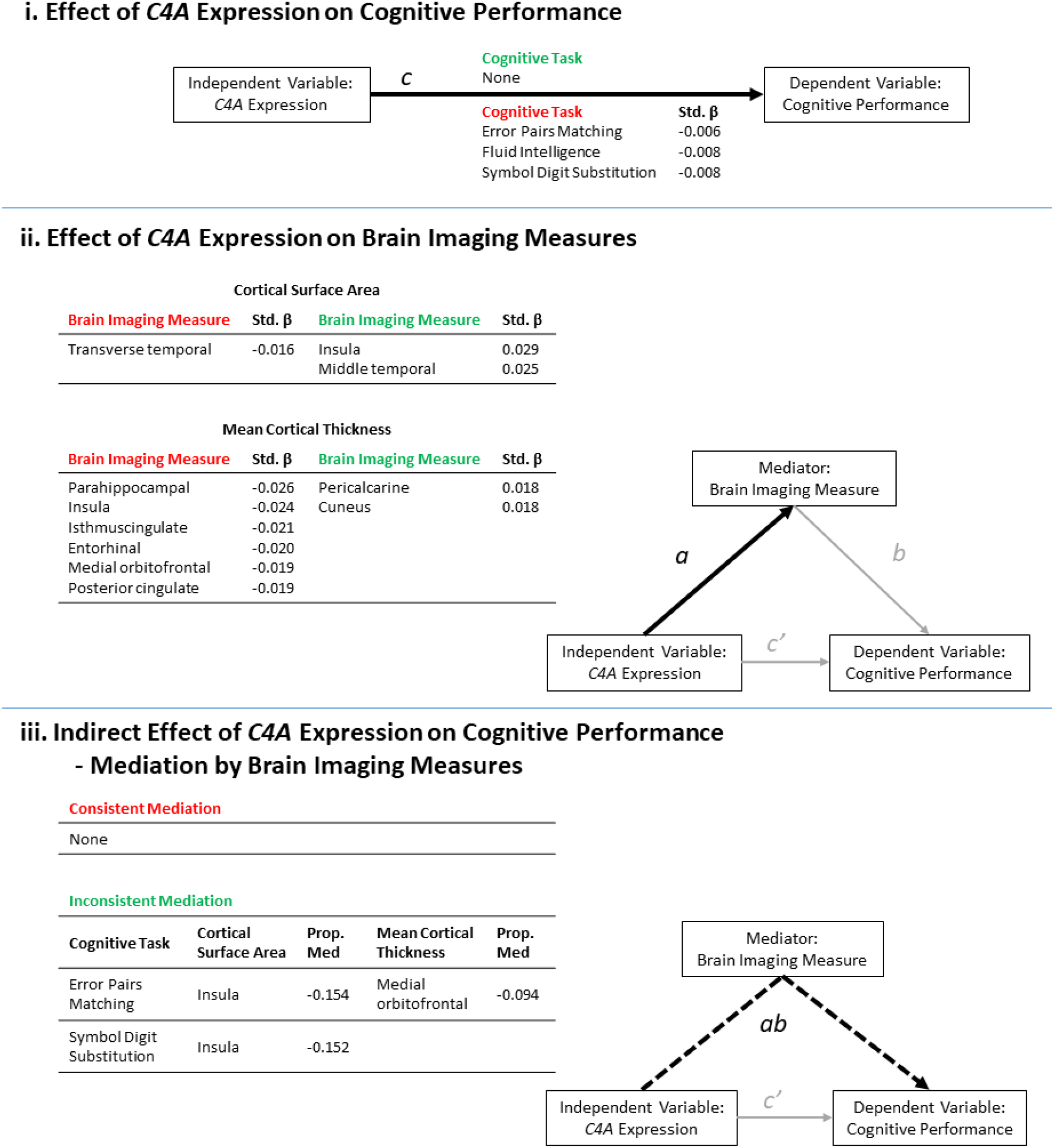
A summary of the results from the significant (FDR < 0.05) linear regression models of predicted *C4A* expression values on cognitive performance and brain imaging measures. The results are presented in the context of a mediation model. (i) Higher predicted *C4A* expression was significantly associated with the results from three cognitive tasks. Path *c* = Cognitive task ∼ *C4A* expression (ii) Predicted *C4A exp*ression was significantly associated with some measures of cortical surface area and cortical thickness. Path *a =* Brain imaging measure ∼ *C4A* expression. (iii) A summary of the brain imaging measures identified to significantly mediate the effect of predicted *C4A exp*ression on cognitive performance. Path *ab* = Cognitive task ∼ *C4A* expression mediated by brain imaging measures. The proportion of the total effect (Panel i, Path *c*) mediated by changes in the corresponding brain imaging measure is shown (Prop. Med = *ab*/*c*). Negative proportion values indicate inconsistent mediation. Inconsistent mediation occurs when the direction of effect of the direct effect (*c’*) and the indirect effect (*ab*) are in the opposite direction. The standardized β (Std. β) shown to indicate the size and direction of effect of higher *C4A* expression on each outcome measure. The green and red headers indicate an increase or decrease in each outcome measure, respectively.

### Effect of *C4A exp*ression on Brain Imaging Measures

Predicted *C4A exp*ression was significantly (FDR < 0.05) associated with three cortical surface area measures (Figure 2A, Supplementary Table 8). Specifically, higher *C4A* expression was associated with increased surface area for the transverse temporal measure (FDR = 0.045), and reduced surface area of the insula (FDR = 1.735 × 10^−4^) and middle temporal (FDR = 7.458 × 10^−4^) measures, respectively (Figure 1ii).

**Figure 2.**
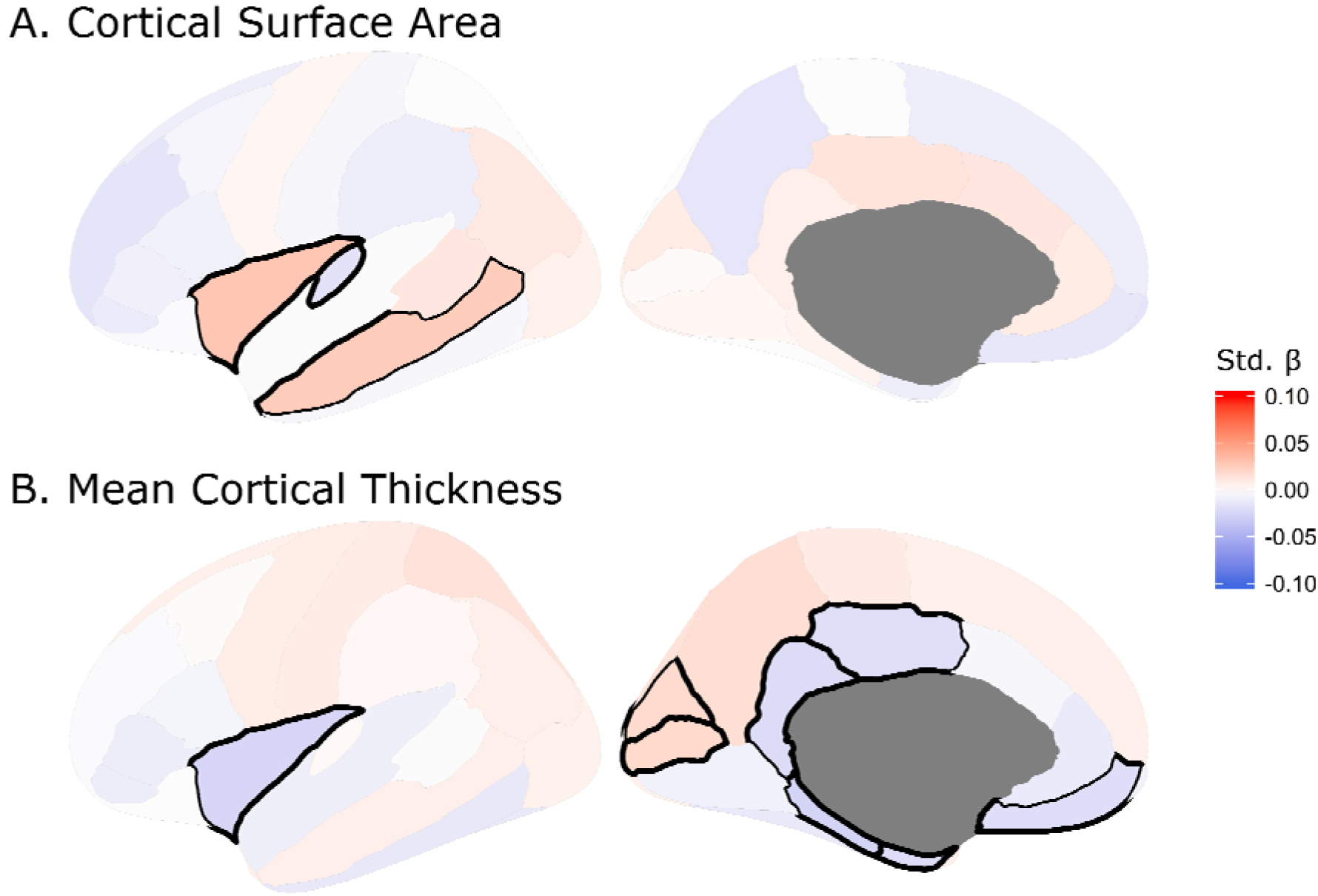
The effect of *C4A exp*ression on regional measures of (A) cortical surface area and (B) mean cortical thickness. The colors correspond to the standardized β (Std. β) coefficient for each brain region from the linear regressions. Black demarcations around a brain region indicates that it passes the multiple comparisons–corrected significance threshold of FDR < with thicker lines indicating more significant findings.

When considering mean cortical thickness, predicted *C4A exp*ression was significantly associated with eight measures, the majority (6 of 8) of which were negatively associated with *C4A* expression (Figure 2B, Supplementary Table 9). Specifically, the parahippocampal (FDR = 7.458 × 10^−4^), insula (FDR = 1.277 × 10^−3^), isthmuscingulate (FDR = 9.865 × 10^−3^), entorhinal (FDR = 0.014), medial orbitofrontal (FDR = 0.016) and posterior cingulate (FDR = 0.017) measures (Figure 1ii).

No significant associations were identified between predicted *C4A* expression and subcortical volumes. In addition, no other regional brain measures, or global measures including total cortical surface area, total mean cortical thickness and ICV, were significantly associated with predicted *C4A exp*ression (Supplementary Tables 8–10). As with cognitive performance, further analysis of the association between predicted *C4A* expression and regional brain imaging measures indicates that this relationship is linear, and that there is not a distinct range of expression above or below which the observed changes occur (Supplementary Tables 10–11 and Supplementary Figures 2–3). Hemisphere-specific results are provided in the supplement (Supplementary Tables 11–13). A summary of the effects of predicted *C4A exp*ression on brain imaging measures, and how these results are incorporated into the mediation analyses is shown in Figure 1ii.

No significant *C4A*-sex interactions were identified for any brain imaging measures (Supplementary Tables 14–16). The same significant associations were identified between higher predicted *C4A* expression and regional measures of cortical surface area and thickness in males only (Supplementary Tables 17–19), but not in females (Supplementary Tables 20–22). With the exception of mean entorhinal thickness which was significant in females only, and not in males.

### Indirect Effect of *C4A* Expression on Cognitive Performance – Mediation by Brain Imaging Measures

Mediation analyses highlighted that increases in insula surface area and medial orbitofrontal thickness are linked to significant (FDR < 0.05) inconsistent mediation of the effect of higher predicted *C4A exp*ression on two measures of cognitive performance (Figure 1iii), i.e. the changes in brain imaging measures partially suppress the negative effects of higher *C4A exp*ression on cognitive performance. None of the included brain imaging measures were identified as significant mediators of the effect of predicted *C4A exp*ression on fluid intelligence scores (Supplementary Table 23).

## DISCUSSION

Here we identified novel significant associations between predicted brain *C4A* expression and cognitive performance in a large adult volunteer sample of individuals without mental or neurological disorders. Additionally, we showed that predicted *C4A* expression was significantly associated with regional cortical thickness and surface area. Further analysis of these associations revealed that their relationships are linear, and that there is no distinct threshold value for predicted *C4A* expression, highlighting that multiple factors likely influence cognition and brain morphology in these individuals within the normal range. Finally, we identified significant inconsistent partial mediation of the effects of *C4A* expression on cognitive performance, by specific brain imaging measures. This indicates that the observed changes in brain morphology may help to protect against *C4A*-associated cognitive deficits. In addition, our observations of changes in cognitive performance and brain imaging measures are highly unlikely to be secondary to any mental or neurological disorders or the treatment thereof. This is because we excluded all those individuals with diagnosed mental or neurological disorders, and the remaining individuals within the UK Biobank tend to be healthier than the general population (Fry et al., 2017).

The main finding of this study is the negative association between predicted *C4A* expression in the brain and episodic memory (Pairs Matching task), reasoning and problem solving (Fluid Intelligence test) and complex processing speed (Symbol Digit Substitution task). Our regression modelling shows that the effects of predicted *C4A* expression are comparable in size to known modifiers of cognitive performance, such as age or sex (Table 1). As expected, when comparing these effect sizes to those of rare copy number variants (CNVs) with known cognitive effects, a study on the same UK Biobank participants showed that most such CNVs had greater effect on cognitive performance than that observed for predicted *C4A* expression in this study (Kendall et al., 2019). These results are in line with previous findings, that higher predicted *C4A* expression is associated with poorer performance in memory recall measures in psychosis patients (Donohoe et al., 2018) and that the complement system modulates memory loss (Wang et al., 2020), and further demonstrate that these effects are present within unaffected individuals. Predicted *C4B* expression was not associated with cognitive performance, the effect of the C4 locus was limited to *C4A* as suggested by previous findings (Donohoe et al., 2018; Sekar et al., 2016).

Cognitive impairments reliably distinguish between schizophrenia patients and healthy controls, with large effect sizes ranging from 0.64 to 1.20 in meta-analyses (Mesholam-Gately et al., 2009). Moreover, similar observations, with smaller effects (effect sizes 0.26 to 0.36), for measures of processing speed, attention and memory have also been identified when comparing first-degree relatives of schizophrenia patients to healthy controls (Hou et al., 2016). At a molecular level, shared common variants that contribute to both schizophrenia risk and cognitive performance have also been identified (Smeland et al., 2019). These studies highlight cognitive impairment as a core heritable feature of schizophrenia (Barch & Ceaser, 2012; Bora et al., 2010; Heinrichs & Zakzanis, 1998), which may manifest in both affected patients and healthy individuals with some genetic burden for the disorder. Cognitive deficits have been associated with poorer functional outcomes regardless of age, sex or chronicity of the disorder (Fett et al., 2011). This lead to the suggestion that common mechanisms might modulate individual differences within these cognitive domains, e.g. related to the structure, function and/or connectivity of prefrontal, parietal, cingulate and insula brain regions (Barch & Ceaser, 2012). Our brain imaging results highlight that *C4A* expression may potentially act as one of the causative factors in such mechanisms.

We identified significant associations between predicted *C4A* expression and cortical surface area and/or mean cortical thickness within temporal, cingulate and insula cortex, amongst others (Figures 1ii and 2). In line with previous observations of structural brain abnormalities in patients with schizophrenia (Cobia et al., 2011; van Haren et al., 2011; Kubota et al., 2011; Assunção Leme et al., 2013; Moberget et al., 2018; Alnæs et al., 2019), and more recent associations between schizophrenia polygenic score and structure in unaffected individuals (Alnæs et al., 2019; Neilson et al., 2019; Westlye et al., 2019), higher predicted *C4A* expression was mostly associated with smaller cortical surface area and lower mean cortical thickness (7 of 11 brain imaging measures, Figure 1ii). These results, together with our findings on cognitive performance, provide further evidence that some of the common genetic underpinnings of schizophrenia may have similar effects also in individuals without mental disorders, in line with dimensional and polygenic risk models (Boyle et al., 2017; International Schizophrenia Consortium et al., 2009; Timpson et al., 2018).

In contrast to these results, higher predicted *C4A* expression was also associated with increased cortical surface area and mean cortical thickness in a subset of brain regions (4 of 11 brain imaging measures, Figure 1ii). Among these regions with an increased cortical surface area are the insula and the middle temporal cortices. This is contrary to what is observed in schizophrenia patients where the cortical surface area of these regions is reduced (Assunção Leme et al., 2013; Cobia et al., 2011; Kubota et al., 2011; van Haren et al., 2011). Interestingly, however, larger cortical surface area has previously been identified in unaffected relatives of schizophrenia patients when compared to non-relative controls (Goghari et al., 2007). That study showed that relatives had increased gray matter volume and surface area in the left hemisphere, bilaterally in the parahippocampal gyri, and in the left middle temporal lobe, thereby implicating the cingulate and temporal regions which are known to be associated with higher level cognitive, affective, and memory functions (Goghari et al., 2007). The authors suggested two possible explanations for these observed increases in the gray matter of relatives; i) abnormal cell migration and deficient pruning, and ii) a protective or compensatory factor against the development of psychosis or loss of associated functioning (Córdova-Palomera et al., 2018; Goghari et al., 2007). Given the molecular functions of complement C4 in the brain, our results could support their suggestion of altered cell migration and synaptic pruning. Moreover, our mediation analyses also suggests the presence of compensatory factors against *C4A*-associated cognitive deficits in individuals without mental disorders.

Previous large scale studies investigating the differences in brain imaging measures between schizophrenia patients and healthy controls show prolific effects of the disorder on numerous measures of cortical surface area and thickness (Theo G. M. van Erp et al., 2018), as well as subcortical volumes (T. G. M. van Erp et al., 2016). Although these effects are considered small to medium (Cohen’s d: 0.2 – 0.5), they are much larger than the effects of *C4A* expression observed in the present study. This highlights that although the changes in brain structure in schizophrenia may be influenced by the level of *C4A* expression, a large number of genetic and environmental factors are likely to contribute, as suggested by previous studies (Lee et al., 2016; Smeland et al., 2018).

Brain imaging measures were shown to correlate positively with general cognitive performance in the UK Biobank (Cox et al., 2019). Since we had identified a significant negative effect of *C4A* expression on cognitive task performance and significant effects on brain imaging measures (predominantly in the negative direction) (Figure 1), we expected *ex ante* to observe consistent mediation via the indirect effect (Figure 1iii, path *ab*), i.e. that some proportion of the effect of *C4A* expression on cognitive performance would be accounted for by the effect of *C4A* expression on brain imaging measures. All of our observations, however, were of inconsistent mediation, i.e. that changes in brain structure, directly or indirectly related to higher *C4A* expression, may act in a protective or compensatory manner against *C4A*-associated cognitive deficits. Significant C4A-associated increases in insula surface area were shown to partially mediate the effects of *C4A* expression on cognitive performance (Figure 1iii). Specifically, increased insula surface area suppressed the negative effects of *C4A exp*ression on episodic memory (Pairs Matching task) and complex processing speed (Symbol Digit Substitution task) by approximately 15% (Figure 1iii). A similar compensatory relationship was identified between *C4A* expression, cognitive performance and mean medial orbitofrontal cortical thickness (Figure 1iii). Increased medial orbitofrontal cortical thickness suppressed the negative effects of *C4A exp*ression on episodic memory (Pairs Matching task) by approximately 9% (Figure 1iii). In this instance, however, predicted *C4A* expression was negatively associated with mean medial orbitofrontal cortical thickness. Therefore, this observed protective effect is likely in response to higher *C4A* expression, and not as a result of higher *C4A* expression.

Partial mediation of the effects of *C4A* expression on cognitive performance, by changes in brain imaging measures, suggests that additional mechanisms play a role in modulating this relationship. Furthermore, given the healthier bias of UK Biobank participants (Fry et al., 2017), further exaggerated by our removal of individuals with mental or neurological disorders, it is tempting to speculate that these participants may share other protective or compensatory factors, in addition to the brain imaging changes identified in this study, which might mask the true effect of *C4A* expression on cognitive performance. Thus, the true effect would likely be greater in an unbiased population cohort. Future studies should identify additional factors associated with changes in *C4A* expression and cognitive performance in order to determine other mechanisms that might contribute to their relationship.

Sex-specific analysis showed that the effects of *C4A* expression on brain morphology were more pronounced in men than in women, in line with recent literature highlighting *C4* sex-specific disease vulnerability (Kamitaki et al., 2020). The known earlier peak in C4 levels in males (20–30 yrs), compared to females (40–50 yrs), may account for this observed difference (Kamitaki et al., 2020; Ritchie, Palomaki, Neveux, & Navolotskaia, 2004; Ritchie, Palomaki, Neveux, Navolotskaia, et al., 2004).

A limitation to the current study is that the UK Biobank has an older age distribution in comparison to patients included in most schizophrenia studies, which are commonly conducted on individuals within an age range more closely matching the age of onset of the disorder (18–25 yrs). As a result, despite controlling for age in our analyses, we cannot exclude a potential effect of aging on the results. Studies in prospective cohorts are required to address this limitation. A second limitation is the reduced sample size for some of the cognitive tasks (n < 100,000). Since the identified significant effects of *C4A* expression of cognitive tasks were small, and predominantly identified for those tasks with the largest sample sizes, these reduced numbers may have resulted in false negatives. Future studies with larger samples for these cognitive tasks are required to determine their true relationship with *C4A* expression.

In conclusion, we observed that higher predicted *C4A* expression is associated with lower cognitive performance and regional cortical surface area and thickness. Moreover, we provide evidence that the observed changes in cognitive performance, as a result of predicted *C4A* expression, may be mediated by C4A-associated changes in brain structure. These results demonstrate that the *C4* locus affects cognition and brain morphology in individuals without mental or neurological disorders.

## METHODS

### The UK Biobank Cohort

The UK Biobank cohort and available data is described elsewhere (Bycroft et al., 2018). Briefly, the UK Biobank project is a prospective cohort study with genetic and phenotypic data collected on approximately 500,000 individuals from across the United Kingdom. Multimodal imaging assessments are underway, with magnetic resonance imaging (MRI) of the brain currently available for a subset of individuals (Miller et al., 2016). All data used in this study were obtained from the UK Biobank (http://www.ukbiobank.ac.uk) through application 27412.

We limited the cohort to 409,629 Caucasian individuals (Datafield-22006). This subset is defined as those individuals who self-identified as ‘White British’ and that had similar genetic ancestry based on a principal component analysis. Individuals with a diagnosed mental or neurological disorder were excluded (Datafields-41202,41204; F/G codes). One from each pair of individuals with a kinship coefficient above 0.053 was also removed prior to analyses (Datafield-22011,22012).

The final cohort sample size, after exclusions, with available genetic data was 329,773 (median age 59, range: 40–74). The sample included 152,966 men (median age 59, range: 40–74) and 176,807 women (median age 58, range: 40–71).

All participants provided informed consent prior to enrolment. The study was approved by the South-Eastern Norway Regional Committees for Medical and Health Research Ethics.

### Genotyping and QC

Genotyping of the UK Biobank cohort was performed on two similar arrays. Approximately 50,000 samples were genotyped on the UK BiLEVE array and the remaining 450,000 samples were genotyped on the UK Biobank Axiom array. Further details regarding genotyping and quality control procedures for the UK Biobank are well documented (Bycroft et al., 2018).

### Imputation of C4 structural variation and genetically predicted C4A expression

Direct genotypes for variants (n = 3213) within the MHC region were used to impute C4 structural variation. This analysis was performed using the 222 haplotype-integrated variant and C4 reference panel (Sekar et al., 2016). The distribution of C4 structural variants was similar to previously described (Supplementary Table 24) (Sekar et al., 2016; Kamitaki et al., 2020). The imputed C4 structural alleles were then used to determine C4 isotype (C4A, C4B, C4L and C4S) copy numbers. Here C4A and C4B refer to the two isotypes of the *C4* gene, while C4L and C4S refer to ‘long’ and ‘short’ forms of the gene due to the presence or absence of a human endogenous retroviral (HERV) insertion, respectively. We calculated values for the predicted expression of the *C4A* gene in human brain tissue, based on the previously identified relationship between C4 isotype copy number and *C4A* gene expression (Sekar et al., 2016). The predicted *C4A* expression values ranged between 0 and 2.35 (mean = 1.08, standard deviation = 0.36) (Supplementary Figure 1). A summary of this methodology is presented in Figure 3. Predicted *C4B* expression values were calculated following a similar approach (Sekar et al., 2016). Predicted *C4A* and *C4B* expression values were used for association with cognitive tasks and brain imaging measures since these variables allow for use of standard linear regression analyses instead of ordinal regression using structural variants.

**Figure 3.**
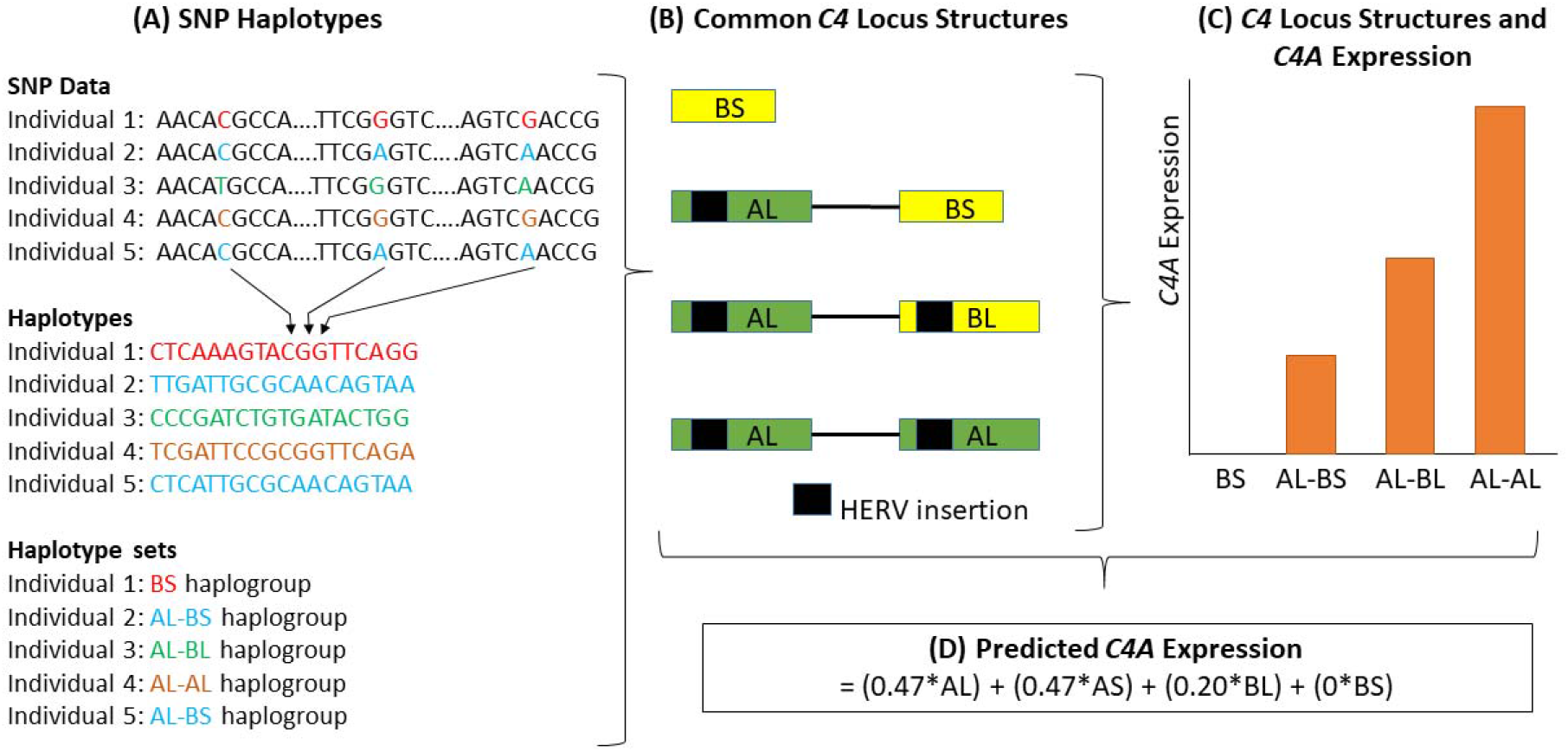
A schematic diagram of the methodology used to obtain predicted expression values for the *C4A* gene within brain tissue, as described by Sekar et al. (Sekar et al., 2016). First, (A) individual genotypes are determined and SNP haplotypes are then inferred from this data. (B) The SNP haplotypes can be grouped into haplogroups and each haplogroup corresponds to a specific *C4* locus structure. Four of these structures are common (represented here) and 11 are rare (< 10% frequency combined). HERV, human endogenous retroviral insertion. C) Structures with higher copy numbers of *C4A* and *C4L* (both *C4AL* and *C4BL*) isotypes show higher *C4A* expression in brain tissue. (D) *C4A* gene expression can be predicted based on the data outlined in panels A-C. AL, AS, BL and BS refer to the copy number of each of these isotypes in the *C4* locus structure. Structures containing the AS combination are omitted from panels A-C since they are rare, with a frequency of approximately 1% (Supplementary Table 24) (Sekar et al., 2016). This figure is a schematic and was not generated from actual genotype, expression or other data.

### Cognitive Tasks

We obtained performance measures on seven cognitive tasks from the UK Biobank, and processed them following a previously described approach (Kendall et al., 2017, 2019). Briefly, measures for analysis included the Pairs Matching task (episodic memory, Datafield-399), the Reaction Time task (simple processing speed, Datafield-20023), the Fluid Intelligence test (reasoning and problem solving, Datafield-20016), the Digit Span task (numeric working memory, Datafield-4282), the Symbol Digit Substitution task (complex processing speed, Datafield-20195), and the Trail Making A and B tasks (visual attention, Datafields-20156,20157). All data was recoded so that higher scores indicate better performance. The number of participants that completed each of these performance measures, with available predicted *C4A* and *C4B exp*ression values and brain imaging data, is provided in Table 2.

**Table 2.** Numbers of participants that completed each of the seven cognitive tasks, with available predicted *C4A* and *C4B* expression values and brain imaging data.

| **Cognitive Task** | **With Predicted *C4A* and *C4B* expression (n)** | **With *C4A* and *C4B* expression Values and Brain Imaging Data (n)** |
| --- | --- | --- |
| Pairs matching | 329,465 | 21,989 |
| Reaction time | 327,815 | 22,064 |
| Fluid Intelligence | 106,633 | 7,484 |
| Digit span | 34,171 | 2,195 |
| Symbol Digit Substitution | 81,444 | 11,696 |
| Trail Making A | 71,933 | 10,427 |
| Trail Making B | 71,931 | 10,427 |

### Image acquisition and processing

Imaging assessments were conducted at three centers, using the same hardware, software and protocols. A detailed description of the processes for data acquisition, processing and quality control is available (Alfaro-Almagro et al., 2018). The data release from UK Biobank used in this study included 33,003 participants. *C4A* and *C4B exp*ression values were predicted for 27,087 of these participants.

We processed T1-weighted MRI scans from all individuals using the standardized recon-all pipeline of FreeSurfer (Fischl et al., 2002; Fischl, 2012). Analyzed brain imaging measures included surface area and mean thickness of 34 cortical regions, total cortical surface area and mean cortical thickness, the volumes of seven sub-cortical regions, and total intracranial volume (ICV). The total surface area, thickness or volume of each region was calculated by summing the right and left hemispheres.

### Statistical Analyses

To determine the relationship between cognitive performance and predicted *C4A* and *C4B* expression, we performed linear regression analyses with each cognitive task as the outcome variable, predicted *C4A* or *C4B* expression as the predictor variable and common covariates, which included age, age-squared, sex, genotyping batch, the first 10 genetic principal components and educational attainment. Age-squared was included since this allows the model to accommodate a non-linear relationship between age and the outcome variable, if one exists. Educational attainment was determined by the highest qualification obtained by each individual at the time of assessment (Datafield-6138). No significant associations were identified between predicted *C4B* expression and cognitive tasks (Supplementary Table 25), and therefore predicted *C4B* expression was not tested for associations with brain imaging measures.

To investigate the relationship between brain imaging measures and predicted *C4A* expression values, brain imaging measures were first normalized in R 3.5.0 by an inverse normal transformation of the residual of a linear regression on the phenotype correcting for covariates, as previously described (Sønderby et al., 2018). This transformation results in normally distributed covariate-corrected values that were used for downstream analysis. Covariates included the common covariates mentioned above as well as Euler number (Rosen et al., 2018). Regional measures of surface area and mean thickness were corrected for total cortical surface area and total mean cortical thickness, respectively. Subcortical volumes were corrected for ICV.

To determine the association between of predicted *C4A* expression and brain structure we performed linear regression analyses with the covariate-corrected brain imaging measure as the outcome, and predicted *C4A exp*ression as the predictor variable in the model.

Finally, to determine if the effects of predicted *C4A exp*ression on cognitive tasks were mediated by brain imaging measures, additional linear regression analyses were performed with each cognitive task as the outcome variable, predicted *C4A exp*ression, a regional noncovariate-corrected brain imaging measure and covariates. Covariates included the common covariates, Euler number (Rosen et al., 2018), and educational attainment. Regional measures were corrected for using the global measures as described above. Mediation analysis was then performed using the R package mediation v4.4.6, using the bootstrapping method and 5000 simulations per test (Writing Committee for the ENIGMA-CNV Working Group et al., 2019). All significant results are also shown in the context of a mediation model (Figure 1). A previous study investigating the effects of brain imaging measures on cognitive performance in the UK Biobank has shown significant positive correlations between all of the brain imaging measures included in this study and increased cognitive performance (Cox et al., 2019). Those results correspond to path *b* in the mediation analyses performed in this study (Figure 1).

Since sex-specific *C4A* risk effects were recently identified (Kamitaki et al., 2020), additional analysis were performed as above with the inclusion of an interaction term between *C4A* expression and sex, as well as in males and in females separately (Supplementary Tables 5–7). The number of male and female participants that completed each of the performance measures, with available predicted *C4A* and *C4B exp*ression values and brain imaging data, is provided in Supplementary Table 25.

The distributions of residuals from all models were examined and determined to be normal indicating that linearity assumptions were not violated. Effect sizes reported are the standardized estimates of beta (β) from the linear regressions. The partial correlation coefficient (r) was computed from the t-statistics for the main cognitive and brain structure analyses (Supplementary Tables 1, 8–10). The distribution of values for significantly associated cognitive performance tests and brain imaging measures were plotted against ‘binned’ predictions of *C4A* expression levels (Supplementary Figures 1–3) and analysis of variance tests and post-hoc Tukey tests were used to determine the differences between these ‘bins’ (Supplementary Tables 2–4). Empirical p-values were converted to False Discovery Rate (FDR) q-values using the R package qvalue v2.14.1. Results were considered significant if FDR < 0.05. Plots were generated using R library ggplot2 v2.2.1 (Wickham, 2009, p. 2) and the R package ggseg v1.5.1.

## Data Availability

This research has been conducted using the UK Biobank Resource (Application number 27412)

https://www.ukbiobank.ac.uk/

## ACKNOWLEDGEMENTS

We gratefully acknowledge support from the Research Council of Norway (grant 273291, 248980, 248778 and 248828 for OAA; grant 249795 for LTW), and the South-Eastern Norway Regional Health Authority (grant 2018094 for SD; grant 2020060 for IES; grant 2017–112 for OAA; grant 2019101 for LTW), the European Research Council under the European Union’s Horizon 2020 research and innovation program (ERC Starting Grant 802998: BRAINMINT for LTW). This research has been conducted using the UK Biobank Resource (Application number 27412).

